# Co-creating community driven solutions and policy asks to address antimicrobial resistance through Responsive Dialogues: A qualitative evaluation from Malawi

**DOI:** 10.64898/2025.12.09.25341950

**Authors:** Henry Sambakunsi, MacWellings Phiri, Thomasena O’Byrne, John Mankhomwa, Raymond Pongolani, Jo Zaremba, Nicholas A Feasey, Eleanor Macpherson, Deborah Nyirenda

## Abstract

Low public awareness and inappropriate antibiotic use contribute to the growing Antimicrobial Resistance (AMR) burden, especially in low- and middle-income countries (LMICs). The aim of Responsive Dialogues is to engage communities and key stakeholders to share information, galvanize action and create local solutions to reduce the burden of AMR. RDs promote collaborative problem-solving and have been identified as a potential tool for engaging diverse stakeholders in addressing AMR. However, RDs were only recently developed, and empirical evidence about their utility and feasibility in LMICs is currently limited. We piloted Responsive Dialogues (RDs) with poultry farmers, government and private pharmacists or prescribers, and male caregivers in Malawi. Using 13 semi-structured interviews and three focus groups analyzed thematically, we assessed feasibility of this participatory approach to influence inclusive local AMR policies and solutions. The participatory nature of RDs promoted inclusive decision-making and facilitated participants’ capacity to co-create locally relevant AMR solutions. Information shared during RDs improved AMR awareness, prompting participants to recognize and reconsider antibiotic practices and co-create solutions including community education on responsible use, local stewardship committees, and integrating AMR education into existing health programs. Implementation barriers varied: farmers and private prescribers expressed concerns about economic losses from reduced antibiotic use, while male caregivers reported difficulties educating others without tangible materials or resources.

Considerations for AMR-RDs scale-up include resource allocation, integration with existing health systems, and further research on long-term impact across different low resource settings. Despite these challenges, this pilot demonstrates that RDs offer a feasible participatory approach for co-creating contextually relevant AMR solutions in resource-limited settings, showing promise for engaging diverse stakeholders though sustained impact requires addressing identified barriers and health system integration.

## Introduction

Antimicrobial resistance (AMR) has emerged as a significant global health threat, with particularly severe implications for low- and middle-income countries (LMICs) (1). The World Health Organization (WHO) has identified AMR as one of the top ten global public health threats facing humanity (1). Current estimates suggest that AMR was directly responsible for 1.27 million deaths globally in 2019, with the highest burden in sub-Saharan Africa (sSA) (2). Projections indicate this could rise to 10 million deaths annually by 2050 under worst-case scenarios if left unchecked (3).

In Malawi, where the health system faces resource constraints and the burden of severe bacterial infections remains high, antimicrobial resistance has significant clinical and economic consequences. Recent prospective cohort studies from Blantyre have demonstrated that third-generation cephalosporin resistance is associated with increased mortality and prolonged hospital stays, as well as substantial individual and population-level economic costs (4,5). Surveillance data from the WHO African Region indicates that sSA experiences the highest global AMR mortality rate at 23.5 deaths per 100,000 population, though country-specific comprehensive data for Malawi remains limited (6). Longitudinal surveillance data from Queen Elizabeth Central Hospital (QECH) in Blantyre demonstrates alarming trends in antimicrobial resistance in bloodstream infection isolates from 1998-2016, with significant increases in resistance to commonly used antimicrobials among both pediatric and adult patients (7). This trend not only threatens individual health outcomes but also potentially affects food security, economic development, and the achievement of the Sustainable Development Goals (SDGs) (8).

The use and misuse of antimicrobials in both human and animal health sectors is widely recognised as a key driver of AMR (9). In Malawi, challenges such as inadequate regulation of antibiotic use, limited diagnostic capabilities, and low public awareness combined with a high burden of severe bacterial infection are thought to contribute to the growing threat of AMR (10,11). A recent review documented similar patterns across sSA, where structural health system challenges compound the AMR crisis (12).

Recognizing the urgency of this issue, the Malawian government established a Ministerial AMR Unit to coordinate the country’s response, culminating in a National Action Plan (13). The successful implementation of this plan is understood to require engagement from key stakeholders, including affected community members, to co-create and implement effective interventions (14).

## Responsive dialogues implementation in Malawi

The authors conducted a public engagement initiative from 2020 to 2023 to facilitate RDs with key stakeholders to co-create solutions and enhance inclusive participatory processes to address AMR in Malawi. The initiative piloted RDs, a toolkit for public engagement initially developed by Wellcome and more recently revised by International Centre for Antimicrobial Resistance Solutions (ICARS) in collaboration with the University of the Western Cape (15). The toolkit, developed through iterative consultation with stakeholders in multiple countries, Malawi and Thailand, uses a participatory approach to bring communities, policy makers, researchers, and civil society groups together to consider and reflect on AMR and the evidence behind AMR. The purpose of this approach is to galvanize action and create local solutions that reduce the burden of AMR. A recent publication from a participating country, Thailand, a very different setting, has demonstrated the RD’s potential for generating locally relevant solutions, though long-term impact remains limited (16).

In Malawi, the RDs were piloted with participants from rural, peri urban and urban areas in Blantyre using three phases: 1) scoping and design; 2) RDs implementation; and 3) synthesizing evidence, establishing impact and dissemination. The scoping and design phase involved understanding the AMR ecosystem in Malawi, mapping out key stakeholders (e.g. policy makers, researchers, local leaders, media personnel, healthcare providers, etc.) and existing AMR initiatives. This was done through consultations and in collaboration with the AMR unit at the Malawi Ministry of Health. Following this process, a stakeholder workshop was convened to set project goals and identify priority participant groups. Three target groups were identified: male and female small-scale farmers using antibiotics to raise animals; male and female private and public antibiotic prescribers operating at community level; and male caregivers (prioritized because they were reported to be less engaged in health research or formal healthcare, potentially more likely to use unprescribed antibiotics, and often responsible for household decision-making).

Implementation of RDs involved what the toolkit calls Conversation Events (CEs). CEs were day-long meetings where the identified stakeholders and participants together explored AMR topics and co-created solutions to address AMR. For each participant group, several CEs were held, and specific stakeholders were invited. The implementation period coincided with the COVID-19 pandemic, requiring adaptations to ensure participant safety while maintaining engagement quality. The third phase focused on collating and synthesizing learnings from the first two phases of work. The overall aim of this phase was to capture solutions, and policy asks that could be presented to influential stakeholders through face-to-face meetings and policy briefs.

RDs are a relatively new approach, with Malawi and Thailand being among the first countries to pilot the toolkit. While community engagement approaches have shown promise in addressing public health challenges(17), there is limited evidence on RDs effectiveness in tackling AMR, particularly in LMICs.This study explored RDs’ role in driving local AMR policies and solutions, contributing to the growing body of knowledge on effective strategies for combating AMR in resource-limited settings. This study presents a qualitative evaluation of the RDs pilot in Malawi, examining how participants experienced the engagement process, what solutions they co-created, and what factors they identified as facilitating or hindering implementation.

## Methods

### Data collection

This qualitative evaluation was conducted between February 2022 and July 2022 approximately 3 months after the final Conversation Events held in November 2021. Data were collected through semi-structured interviews (n=15) and focus group discussions (FGDs) (n=3) to capture both individual perspectives and group dynamics in participants’ experiences of the RDs process. SSIs allowed for in-depth exploration of personal experiences and changes in practice, while FGDs facilitated discussion of shared challenges and collective reflections on implementation barriers. Each FGD had eight participants with equal gender representation (see Table 1), lasting approximately 90-120 minutes.

**Table 1:**
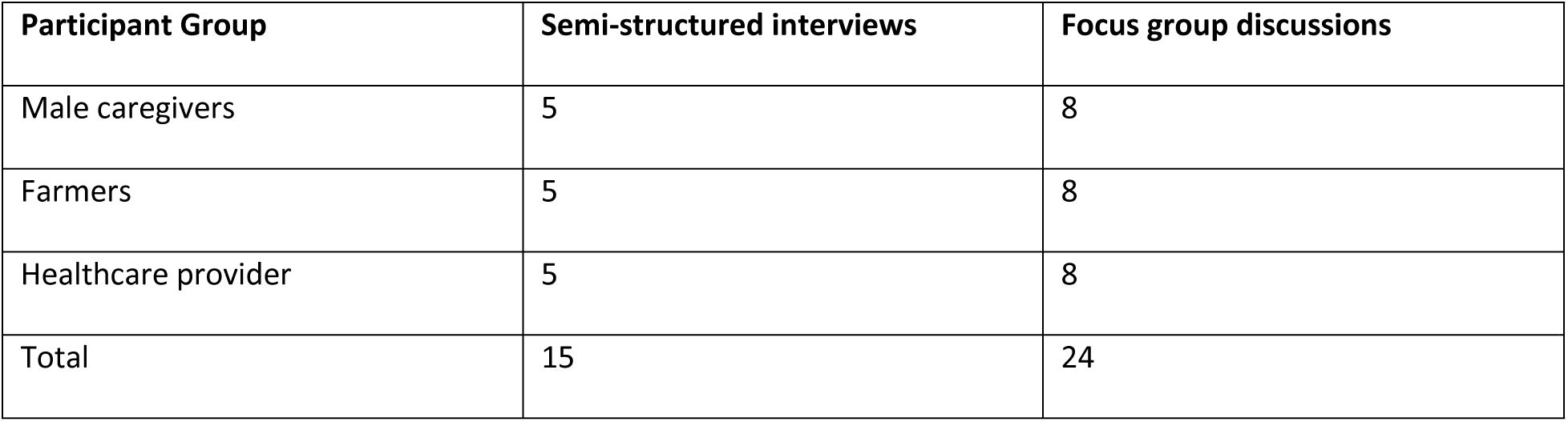
Participants for semi-structured interviews and focus group discussions.

A topic guide was developed to facilitate discussion about stakeholder’s experiences as co-producers of AMR solutions and feasibility of co-created solutions. Broad topics of the guide included perceptions about the RDs toolkit, views on the process of implementing the RDs, experiences of participating in the project including perceived successes and challenges of implementing the co-created solutions. We also asked the participants to identify self-reported changes in knowledge and practices, discuss perceived barriers and enablers to implementing solutions, and participants views on whether the co-produced solutions might be sustained beyond the workshops. These topics were formulated by the research team when developing the research protocol and were iteratively modified throughout data collection. All the interviews and FGDs were conducted in Chichewa by a native-speaking interviewer (HS) and with support for note taking from (RP).

### Participants

The study participants included individuals from the three main groups of participants in the Conversation events namely livestock farmers (cattle and poultry), male caregivers from the community, and healthcare workers including nurses, pharmacists, and clinicians who participated in the Responsive Dialogues workshops.

Participants were purposively selected from Responsive Dialogues workshop attendees to ensure representation across all three participant groups (farmers, healthcare providers, male caregivers) and geographic settings (rural, peri-urban, urban areas in Blantyre). Potential participants were identified from workshop records and invited to participate by the research team based on their availability and willingness to discuss their experiences. Following identification and contact by the research team, participants were provided with the information sheet and consent form and given the opportunity to ask further questions about the study. Participants were reassured that their involvement was voluntary, their contributions would be kept confidential, and participation would not impact their involvement in future workshops or services. The interviews and FGDs were undertaken at a time and place convenient to the participant.

### Ethics statement

Human research ethical approval was received through the College of Medicine Research Ethics Committee (COMREC), in Malawi (Protocol Number: P.07/21/3360) on 15 November 2021, prior to commencement of the research. The study was conducted in accordance with the ethical principles outlined in the Declaration of Helsinki.

Participant recruitment for this qualitative evaluation took place between February 2022 and July 2022. Written informed consent was obtained from all participants prior to their participation in semi-structured interviews and focus group discussions. Participants were provided with information sheets in both English and Chichewa explaining the study purpose, procedures, potential risks and benefits, confidentiality measures, and their right to withdraw at any time without consequences. All participants were adults aged 18 years and above who had previously participated in the Responsive Dialogues Conversation Events. Participants were reassured that their involvement was voluntary, their contributions would be kept confidential, and participation would not impact their involvement in future workshops or services.

### Data management and analysis

All IDIs and FGDs were audio-recorded, transcribed verbatim, and translated into English. The translated transcripts were randomly checked to ensure data accuracy and analysed using inductive and deductive thematic analysis approaches. The steps used included initial and ongoing familiarisation with data, development of codes using NVivo 12 Software, including indexing and charting, summarising/interpretation of data, and the development of themes.

To ensure accurate interpretation of the data, quality control measures were put in place. Data were systematically coded by HS, initial coding was reviewed by DN, and developed codes and themes were discussed and further refined by team members MP, JM, RP, EM. Once the preliminary analysis was completed, consultation sessions were held with all researchers.

Quotes are presented to illustrate the findings, with abbreviated identifiers provided for each participant group (HCP = healthcare provider; FC = farmer/community member, MCG= Male Care giver).

## Results

Evaluation of our implementation of the Responsive Dialogues approach in Malawi revealed four main themes: 1) Improved participant understanding of AMR, 2) Change in attitudes towards antibiotic use and AMR, 3) Challenges impacting implementation of co-created solutions, and 4) Feasibility and limitations of the RD process.

### Improved participants’ understanding of antimicrobial resistance

Data from both focus group discussions and key informant interviews emphasized the perceived transformative role of RDs in disseminating information and improving people’s knowledge regarding AMR. Most participants reported that they knew little or nothing about AMR before the RDs. The qualitative assessment showed how involvement in RDs appeared to transform understanding across the various participant groups.

Male caregivers, who initially reported limited knowledge of AMR, gained awareness through the workshops. Participants characterized the dialogues as an eye-opener, with many encountering the term "antimicrobial resistance" and its public health implications for the first time. The interactive nature of the dialogues allowed participants to engage with the information actively, ask questions, relate the concepts to their own experiences, and share new ideas with their peers. This participatory approach appeared to facilitate a deeper understanding of AMR, moving beyond mere factual recall to what participants described as a more comprehensive understanding of the issue. Caregivers reported coming to recognize the dangers associated with AMR and the importance of correct antibiotic use. The realization that common practices, such as self-medication or not completing prescribed courses of antibiotics, could contribute to this global health threat was described as eye-opening. This reported newfound awareness extended beyond the immediate health implications, encompassing an understanding of the broader societal and economic impacts of AMR.

> "At first, I didn’t know about Antimicrobial Resistance, but after participating in the conversation events that we had with our colleagues, that’s when I gained knowledge that antimicrobial resistance is dangerous and that using drugs without following proper instructions is a bad habit." (MCG, SSI 014)

Similarly, farmers’ self-reported understanding of antibiotics and AMR improved substantially, marking what appeared to be a significant shift in knowledge within this crucial stakeholder group. Prior to their participation in the Responsive Dialogues, most farmers reported having limited or incorrect information about antibiotics, their uses, and the potential consequences of their misuse. The events were described as serving as a transformative educational experience, introducing farmers to the concept of antibiotics as specific medications designed to combat bacterial infections. This reported newfound knowledge extended to an understanding of AMR and its implications for both animal husbandry and human health.

> "I also want to add that when I was coming to participate in those events, I never knew what an antibiotic was, but after participating in the events, that’s when I gained some knowledge on what an antibiotic is and what it does. To me, it was an eye-opener, and we shared new ideas with our fellow farmers." (Participant 3, FC, FGD) Healthcare providers, despite their professional background, also reported enhanced understanding, highlighting the potential impact of the RDs process. This revelation is particularly significant given that these individuals already possessed a foundational understanding of antibiotics and their use through their formal medical training. The fact that even healthcare professionals reported enhancing their comprehension of AMR underscores the complexity and evolving nature of this global health challenge. For most, their previous exposure to AMR concepts in academic settings had not fully conveyed the scale and urgency of the issue. The dialogues served to bridge the gap between theoretical knowledge and practical understanding, providing a more comprehensive and up-to-date perspective on the AMR crisis.

> "I heard about it from college in the Pharmacology course, but I never knew that the problem is that huge until I participated in the meetings…" (HCP, SSI 07)

### Reported change in attitudes towards antimicrobial resistance and antibiotics use

Following participation in the Responsive Dialogues, both FGD and SSI participants’ reported intentions to use antibiotics responsibly. Most participants, especially male caregivers and farmers stated that the RDs helped them to become more cautious about antimicrobials and expressed intentions to refrain from using them without a prescription. This reported shift in attitude may represent a significant outcome of the RDs, suggesting that improving understanding of AMR could potentially lead to more responsible antibiotic use practices at the community level.

> "Before, I used to think that antibiotics were a cure for everything. But after participating in the RDs and learning about AMR, I realized the importance of using antibiotics only when necessary and as prescribed. I’ve changed my attitude towards antibiotic use and I’m more cautious now." (MCG, SSI 014)

Public healthcare workers who prescribed antibiotics at the community level reported similar behavioural change, describing adjusting their drug management practices. Most of them reported a decline in their tendency to collect and keep antibiotics at home for self-prescription. They expressed that they now preferred professionally delivered prescription because they reported better understanding of the risks of antibiotic resistance and the importance of proper medical oversight in antibiotic use. This reported change in behaviour among healthcare professionals is noteworthy, as it suggests that the RDs may have been effective in influencing not only community members but also those who play a crucial role in antibiotic stewardship.

> "I will do differently especially at home, because I had a bad habit of taking drugs here and store them at home so that if my child gets sick when I’m at work he can take the medicine at home, but now I make sure that he gets prescription first." (HCP, SSI 06)

Participants, including most of the male caregivers, also expressed intentions to promote responsible antibiotic use more widely in their communities. Many reported a sense of responsibility to educate their friends, family, and neighbours about the proper use of antibiotics and the risks of AMR. They described plans to organize community meetings, distribute informational materials, and engage in one-on-one conversations to share their knowledge.

Some participants even expressed interest in collaborating with local health facilities and schools to reach a broader audience. This reported desire to improve awareness appears to stem from their realization that AMR directly impacts their own health outcomes and those of their families and neighbors, combined with recognition of the critical role that individual actions play in combating it.By taking on this advocacy role, participants hoped to create a ripple effect of positive change, fostering a community-wide culture of responsible antibiotic use that could contribute to preserving the effectiveness of these crucial medications.

> "I feel a responsibility to educate others about AMR and the proper use of antibiotics. I want to organize community meetings and share the information I learned in the RDs. Together, we can make a difference." (Participant 3, FC, FGD)

### Feasibility and limitations of the Responsive Dialogues process

Both FGD and SSI participants reported viewing the RDs process positively, expressing appreciation for the inclusive discussions and competent facilitation. Many participants reported that the inclusive nature of the RDs appeared to allow for a diverse range of perspectives to be heard. They stated that the facilitators seemed to demonstrate a high level of expertise in guiding the conversations, reportedly ensuring that complex topics were broken down into understandable components. Participants described that the depth of information provided, particularly regarding the mechanisms of antibiotic action and the distinction between antibiotics and other medications such as painkillers, was valuable. They reported that the interactive format of the RDs appeared to encourage active participation, seemingly allowing attendees to ask questions, share their own experiences, and engage in meaningful dialogue with both experts and peers.

> "Our interaction was very good and especially on these issues of antibiotics they really went deep on it… They were giving us insights on how the antibiotics work and how painkillers work." (HCP, SSI 07)

The final co-creation event was reported as being particularly well-received by participants. Many described this event as a highlight, bringing together diverse stakeholders from various backgrounds, all united by their shared interest in addressing AMR. Participants reported that the convergence of different perspectives including from healthcare professionals and community leaders to policymakers and researchers, appeared to create a dynamic environment. They reported appreciating the opportunity to engage in intersectoral policy dialogue during this event.

> "The final event was an outstanding event because it brought various people together whereby, we shared ideas and co-created ideas which we thought would be helpful to the whole country." (MCG, SSI 011)

However, participants also reported several limitations and areas for improvement in the RD process. Many participants expressed that more time was needed for discussions, highlighting what they perceived as a significant constraint in the otherwise well-received approach. They reported that this perceived time limitation reflected the complexity and depth of the topics being addressed, particularly in relation to AMR and its multifaceted implications. Participants stated that they found themselves grappling with numerous interrelated issues, from the scientific aspects of AMR to the society and economic factors influencing antibiotic use. They reported that the time constraints often led to a sense of rushing through important topics, potentially curtailing deeper exploration, and thorough understanding.

> "The only problem that I encountered was time, we were not having enough time. I’m saying this because we were having several topics to discuss so sometimes due to lack of time we were being rushed to finish one topic." (MCG, SSI 013)

### Challenges impacting implementation of co-created solutions

Through the RD process, participants co-created several context-specific solutions to address AMR in their communities. These included community-led awareness campaigns on responsible antibiotic use, local antibiotic stewardship committees, integrating AMR education into existing community health programs and promoting alternative medicine to reduce antibiotic use in livestock. To facilitate community dissemination of these messages, a local cartoonist translated the co-created solutions into culturally appropriate visual materials depicting common scenarios of antibiotic misuse and proper use practices (Figure 1). These cartoon-style illustrations portrayed locally relevant situations such as seeking antibiotics without prescription, sharing medications, and consulting healthcare providers for proper diagnosis, making the AMR concepts accessible to community members with varying literacy levels.

> "The ideas we came up with together, like the awareness campaigns, are things we can start doing in our communities right away. It gives us a way to make a difference." (Participant 2, MCG, FGD)

**Figure 1:**
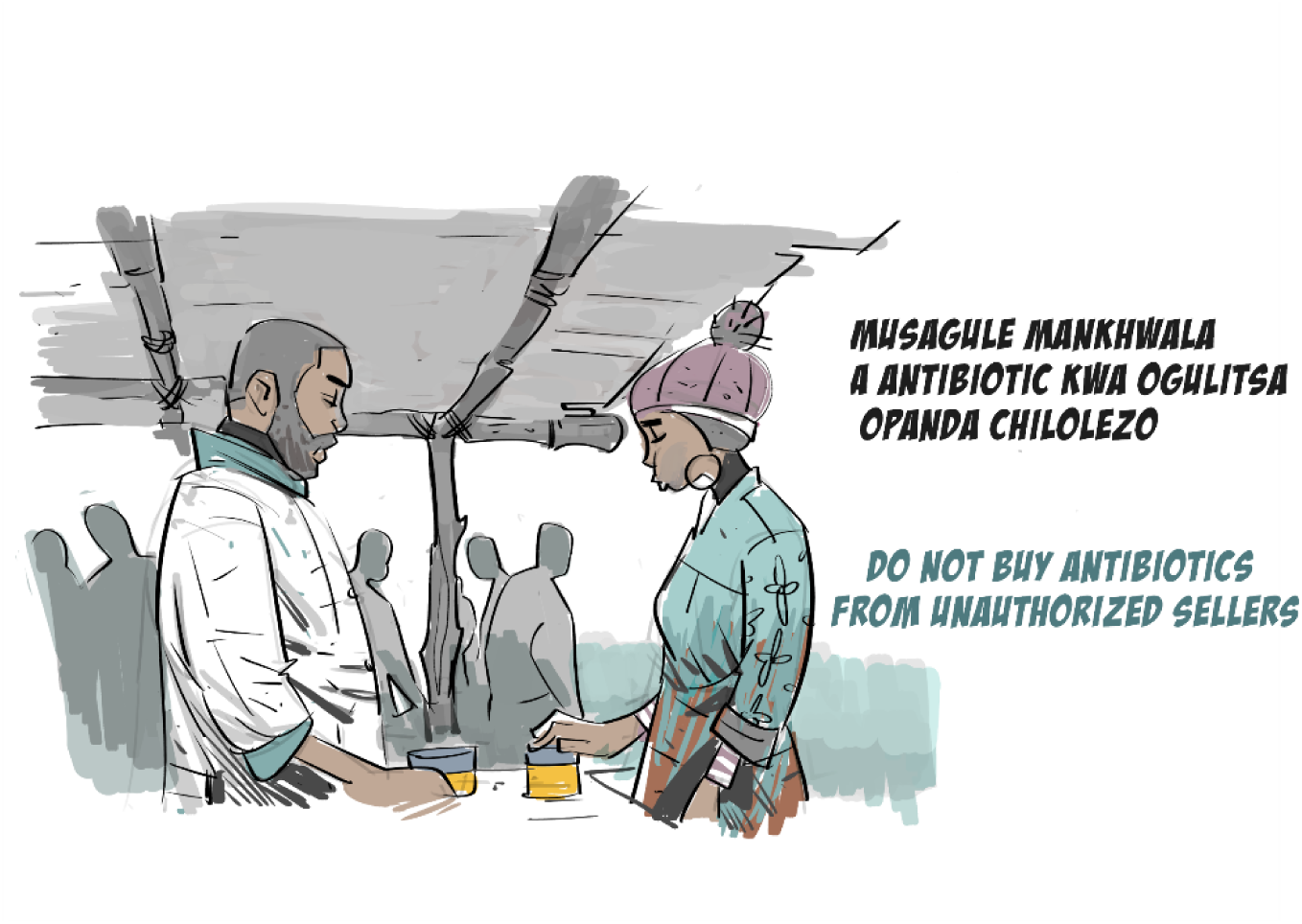
Co-created visual education materials from Responsive Dialogues in Malawi FGD and SSI participants however expressed mixed views about these co-created solutions. Some reported being enthusiastic about local action, seeing the proposed initiatives as tangible ways to make a difference in their communities. These individuals expressed excitement about the prospect of implementing grassroots campaigns and committees, viewing them as potentially empowering opportunities to take control of a global issue at a local level. They reported appreciating how the solutions were tailored to their specific context, making them seem achievable and relevant. The enthusiastic participants were particularly drawn to the immediate applicability of certain ideas, such as awareness campaigns, which they felt could be launched quickly and with minimal resources. Their optimism was fuelled by a sense of collective responsibility and the belief that community-driven efforts could lead to meaningful change in antibiotic use practices.

Despite their reported interest in implementing the co-created solutions to address AMR, many participants also emphasized the need for external support and regulation. While acknowledging the importance of community-driven initiatives, these individuals expressed recognition of the limitations of local action alone in addressing the complex issue of AMR. They argued that comprehensive change would require intervention from higher levels of authority, including government bodies and regulatory agencies. Farmers highlighted the need for stricter enforcement of existing regulations on antibiotic sales, as well as the implementation of new policies to control antibiotic use in both human health and agriculture. They argued that without a robust regulatory framework, community efforts might be undermined by continued easy access to antibiotics through unauthorized channels. Additionally, some participants called for increased financial and logistical support from external sources, such as national health departments or international organizations, to bolster community-driven AMR initiatives.

> "The challenges would be there, because if this behaviour of selling drugs in unauthorized groceries continues then this problem won’t end. We need strict regulations to control the sale and use of antibiotics. People need to understand the consequences of misuse and overuse." (Participant 8, FC, FGD)

Notably, some participants pointed out that the absence of large-scale poultry farmers in the RDs was a significant limitation, revealing a potential gap in the stakeholder engagement process. Participants suggested that the exclusion of large-scale poultry farmers from the discussions represented a missed opportunity to address one of the major contributors to antibiotic misuse in the agricultural sector. This observation highlights the complex nature of AMR, emphasizing the need for an inclusive approach targeting all relevant actors in the antibiotic use chain. Participants stated that the practices of these large-scale poultry farmers could have substantial impacts on AMR development, potentially undermining the efforts made by smaller-scale farmers and other community members. The absence of these key stakeholders meant that their perspectives, challenges, and potential solutions were not integrated into the outputs, thereby limiting the effectiveness of the co-created solutions.

> "However, some people are still misusing antibiotics and most of them are big chicken companies, I’m not sure if they are aware of this issue of antimicrobial resistance because they are still into that bad habit." (Participant 2, HCP, FGD)

Some participants expressed frustration with the lack of feedback on implementation progress. This sentiment appeared to reflect a growing sense of disappointment among those who had invested time and energy in the co-creation process. Many reported feeling that their efforts and enthusiasm had been met with silence or lack of support, leaving them uncertain about the impact of their contributions. The absence of follow-up communication created a perceived gap between the RD’s and any tangible outcomes, leading to questions about the long-term commitment to the initiatives they had co-created. This lack of feedback not only dampened morale but also hindered participants’ ability to gauge the feasibility of the proposed solutions or share experiences based on real-world implementation challenges. For some, this communication void undermined their confidence in RD’s and raised concerns about the feasibility of the AMR initiatives. The frustration expressed by these participants highlighted the critical importance of maintaining ongoing dialogue with community members to achieve long-term public health benefits.

> "I’m saying we were left behind because ever since we finished with the co-creation event there has never been any communication back to answer until now." (Participant 3, FGD)

Participants also highlighted resource constraints as a barrier to implementing the co-created solutions. This concern appeared to underscore the practical challenges faced by community members in translating their newfound knowledge and enthusiasm into tangible action. Many participants expressed frustrations because they believed they possessed valuable information about AMR but lacked the means to effectively disseminate it. The absence of educational materials, visual aids, or other tangible resources made it difficult for them to substantiate their claims and convince others of the importance of responsible antibiotic use. This resource gap not only hampered their ability to conduct awareness campaigns but also undermined their credibility within the community. Some participants felt that without concrete or professional-looking materials, their messages were often met with scepticism or indifference. The lack of financial support for organizing events, printing materials, or even compensating volunteers’ time further compounded these difficulties.

> "For us we have learnt about this issue, but it is becoming difficult for people to believe us when we are sharing the messages with them because we have nothing to show them as evidence of what we are talking about, so it is becoming difficult to influence the change due to lack of the resources." (Participant 5, MCG, FGD)

## Discussion

This study explored the feasibility of RDs to co-create AMR solutions in Malawi. While participants identified numerous benefits of the RDs approach, many also raised concerns related to implementation challenges, resource limitations, and they highlighted the need for sustained engagement.

The global threat of AMR is widely understood to demand efforts to increase public awareness and promote responsible antibiotic use across all sectors (18,19). Concerns however still exists regarding the lack of public awareness about AMR, and the need for context-specific, culturally appropriate interventions (20). Our findings suggest that participants across all stakeholder groups had limited prior knowledge of AMR and antibiotics consistent with broader patterns documented across sub-Saharan Africa (21), including among small-scale farmers in Blantyre where knowledge gaps about antibiotic use and AMR have been documented (22). The RDs approach appeared to address this knowledge gap by providing a platform for knowledge exchange and collaborative learning that participants reported as enabling them to contribute to the co-production of AMR solutions. RDs therefore offer an innovative approach to engaging diverse stakeholders in co-creating solutions to address AMR (18). These findings align with findings from previous studies on participatory approaches suggesting that RDs can potentially increase AMR knowledge, catalyse context relevant solutions to address AMR and reported behaviour changes (23,24). The Thailand RDs pilot reported similar outcomes, including improved AMR understanding among participants with limited prior knowledge, co-creation of context-specific solutions emphasizing local leadership, and resource-intensive implementation challenges. Notably, Thailand documented spontaneous community-led AMR initiatives following the RDs, suggesting potential for sustained engagement beyond the formal project period. However, as in Malawi, longitudinal impact assessments remain limited, representing a critical knowledge gap for both settings (16).

The WHO Global Action Plan on AMR recommends that addressing AMR requires diverse stakeholders to collaborate closely [8]. A key success in the implementation of RDs in Malawi was the inclusive nature of the approach, bringing together healthcare providers, farmers, and community members to co-create solutions. Participants reported appreciating the value of diverse perspectives in co-creating solutions and expressed feeling empowered to contribute to address AMR. This aligns with other studies highlighting the importance of community engagement to empower communities to co-develop sustainable health interventions (25). RD participants also expressed recognition of the need for multi-sectoral collaboration to address AMR, and some reported initial small-scale actions to promote responsible antibiotic use following their participation. The need for sustained action to sustain behaviour changes was a key theme among participants in our study, who expressed seeing the opportunity to build on the momentum created by RDs and integrate some of the innovations into routine practices.

Despite participants expressed willingness to implement small-scale co-created interventions at individual level, our findings highlighted several perceived barriers to implementing AMR solutions, including weak regulations, lack of resources, and challenges in maintaining sustained engagement with communities. This echoes findings from other LMICs, where resource constraints often hinder health intervention implementation (26,27).

The participatory nature of RDs appeared to allow for the co-creation of multi-sectoral interventions, but participants expressed recognition of their limitations in implementing the co-created solutions or influencing meaningful change. To ensure translation and sustainability of the co-created interventions, participants emphasized the importance of strengthening health systems, regulatory frameworks for antibiotics use, financial support for awareness campaigns, and implementation of other interventions. As such, participants suggested that there was need for policy makers to enforce and support some of the co-created solutions about strengthening health systems and drug regulatory frameworks. This is in line with a policy analysis for developing countries that suggests that while National Action Plans exist, implementation gaps remain substantial due to resource and capacity constraints (28). In addition, participants also expressed challenges in translating knowledge into sustained behaviour change even at the individual level. Poverty was highlighted as a significant barrier to reducing antibiotic use in livestock production or limiting sales of antibiotics at community level because most participants reported fearing economic losses. Participants appeared to perceive AMR as a distant threat compared to loss of livelihood and income. This aligns with other studies on behaviour change interventions, which highlight the complex interplay of individual, social, and environmental factors influencing health behaviours (29,30).

The participants in in Malawi expressed a desire for feedback and continued engagement following the RDs. This preference for ongoing communication and support has also been highlighted in other studies on community-based interventions, emphasizing the importance of sustained engagement for long-term impact (31). While RDs appeared to be effective in increasing AMR awareness to co-create solutions and inspire reported behaviour changes, sustained engagement appears to be important to ensure long-term impact. To ensure the sustainability of community-driven AMR solutions, long-term stakeholder support is essential to maintain momentum and foster lasting behavior change. This could include local pilot projects to demonstrate the feasibility and impact of co-created solutions, providing valuable insights and data to inform larger-scale implementation. Future research should also investigate the sustained impact of RDs on antibiotic use practices; identify community-led strategies to fundraise and implement co-created solutions; and investigate how to effectively integrate RDs in other national AMR stewardship initiatives.

### Limitations

The findings of this study contribute to the growing body of evidence regarding participatory approaches to addressing AMR. However, several limitations should be considered when interpreting the results. The main limitation of this study was the focus on short-term outcomes immediately following the RDs workshops. While the findings highlight reported changes in attitudes and behaviours, the long-term sustainability of these changes remains uncertain as previously indicated. Furthermore, the evaluation focused exclusively on participant perspectives and did not systematically include the research team, facilitators, or other stakeholders involved in implementation. Multi-stakeholder evaluation would have provided more comprehensive insights into facilitation challenges, implementation barriers, and the feasibility of scaling up the approach. In addition, self-reported data is inherently subject to social desirability bias, particularly when discussing health behaviors, and should be interpreted with appropriate caution (32). Follow-up studies tracking participants’ sustained engagement with the co-created solutions would provide a more comprehensive understanding of the long-term impact of the RDs approach. Additionally, the absence of baseline data on participants’ antibiotic use practices limits our ability to track actual behavior change. The co-created solutions were intentionally shaped by Malawi’s unique socio-economic context to ensure local relevance. While the participatory RDs approach itself is transferable, the specific solutions would require adaptation when applied in different socio-economic contexts, including other regions within Malawi.

## Conclusion

This study provides valuable insights into the potential of RDs as an approach to engage diverse stakeholders in addressing AMR and shaping public-driven policies and actions, providing rich contextual understanding of implementation challenges and facilitators that can inform future interventions in Malawi. We provide further evidence RDs can effectively support engagement of diverse stakeholders in co-creating multi-sectoral interventions aimed to address AMR. Sustained engagement with community is however required to promote the sustainability and long-term impact of the co-created solutions. In addition, community members and local leaders need to be empowered and supported to effectively implement AMR stewardship efforts. Considerations for scale-up include ensuring equity of access to resources for implementing AMR solutions, fostering policy maker engagement, and integrating RDs outputs with existing health system structures. Future applications should consider integrating outputs from RDs in policy formulation to ensure a comprehensive and sustainable strategy for combating AMR and other global health challenges.

## Data Availability

The qualitative data (audio transcripts) underlying this study contain potentially identifiable information about study participants. The ethical approval granted by College of Medicine Research and Ethics Committee (COMREC) (P.07/21/3360) and the informed consent obtained from participants did not include explicit consent for public sharing of transcripts or anonymized data via a public repository. In accordance with these ethical restrictions and to protect participant confidentiality, the data cannot be made publicly available. Relevant data excerpts supporting the findings are included within the manuscript and supporting information files. Data access requests can be directed to

## Acknowledgements

We are grateful to all participants; poultry farmers, government and private pharmacists and prescribers, and male caregivers who generously shared their time, experiences, and insights during the Responsive Dialogues and subsequent evaluation. We extend our thanks to community leaders in areas of Chemusa, Ndirande, Chileka, Chilomoni in Blantyre for facilitating access and supporting the implementation of this work. We are grateful to the Malawi-Liverpool-Wellcome Programme, Ministry of Health, Ministry of Agriculture and Community Health Science Unit (CHISU) for institutional support.

